# Using Generative AI to Simulate Patient History-Taking in a Problem-Based Learning Tutorial: A Mixed-Methods Study

**DOI:** 10.1101/2024.05.02.24306753

**Authors:** Allison Mool, Jacob Schmid, Thomas Johnston, William Thomas, Emma Fenner, Kevin Lu, Raya Gandhi, Adam Western, Brendan Seabold, Kodi Smith, Zachary Patterson, Hannah Feldt, Daniel Vollmer, Roshan Nallaveettil, Anthony Fanelli, Logan Schmillen, Shelley Tischkau, Anna T. Cianciolo, Pinckney Benedict, Richard Selinfreund

**Author notes:** Correspondence should be addressed to Richard Selinfreund, Department of Medical Microbiology, Immunology and Cell Biology, Southern Illinois University School of Medicine, PO Box 19626, Springfield, IL 62794-9626.

## Abstract

**Background:** Medical educators who implement problem-based learning (PBL) strive to balance realism and feasibility when simulating patient cases, aiming to stimulate collaborative group discussion, engage students’ clinical reasoning, motivate self-directed learning, and promote the development of actionable scientific understanding. Recent advances in generative artificial intelligence (AI) offer exciting new potential for patient simulation in PBL

**Method:** This study used a between-groups, mixed-methods approach to (1) form a comprehensive picture of Year 2 medical student interactions with a generative AI-simulated patient in a PBL tutorial, as compared to interactions with multimedia patient case materials; and (2) triangulate on the impact these interactions had on learning. Two groups of students (N = 13) gathered patient history information from a generative AI-enabled, 3D-animated avatar (AI condition). Two other student groups (N = 13) gathered patient history information from a multimedia database using keyword searching (Electronic PBL Module [ePBLM] condition). We used descriptive observation to explore student interactions with both forms of the simulated patient, and we quantitatively compared students’ perceptions of their learning experience and recall of patient history information across conditions.

**Results:** Students in the AI condition rated their present, AI-augmented PBL learning experience—particularly its clinical accuracy and teamwork aspects—significantly higher than they rated their previous PBL learning experiences using ePBLMs. Recall of patient history information did not differ between conditions. Descriptive observation indicated that the AI avatar presented case content accurately, with an appropriate amount of information provided in response to students’ questions. Students were highly engaged as a group in taking a history from the avatar. However, although students used language suggestive of anthropomorphizing of the AI (e.g., gender pronouns), they appeared to orient to it as an augmented “question bank” for gathering patient history information, using a questioning strategy akin to querying an ePBLM.

**Discussion:** Optimizing AI implementation to stimulate clinical reasoning and patient communication skills in PBL could include (1) starting early, perhaps in Year 1, before an alternative interactional framework can take hold; (2) orienting students to the AI to help them understand its capabilities; and (3) encouraging “play” with or “discovery learning” of the AI’s capabilities.

## Introduction

In medical problem-based learning (PBL), patient cases provide the organizing structure of the curriculum, serving as triggers for learning and providing a platform to exercise thinking representative of physicians’ workplace cognition (Wee, Kek, & Sim, 2001).

Patient case design therefore is a key concern for PBL educators. Quality patient cases stimulate collaborative group discussion, engage clinical reasoning, motivate self-directed learning, and promote the development of actionable scientific understanding.

- Characteristics of a quality patient case – realistic, ideally based on a real patient, set in a medical context, sufficiently challenging for students to stimulate high-level cognitive engagement, designed based on clear learning objectives (Wee, Kek, & Sim, 2001; Azer, Peterson, Guerrero, & Edgren, 2012)
- Characteristics of quality interactions with patient case materials (questioning oneself and others, self-explanation and elaboration of ideas, testing one’s understanding, and self-identification of knowledge and comprehension gaps – Cianciolo & Regehr, 2019)

The diversity of approaches to simulating patients in PBL ranges from paper cases to interactive courseware to live patient actors, each with their own tradeoffs.

- Advantages of high-fidelity simulation (engaging and provides opportunity to practice clinical skills – Barrows & Tamblyn, 1980; facilitates patient-centered learning – MacLeod, 2011)
- Disadvantages of high-fidelity simulation (lack of control and potential to become too complex – Barrows & Tamblyn, 1980; cost in time, personnel, and funding)

Recent advances in generative artificial intelligence (AI) offer exciting new potential for patient simulation in PBL.

- Capabilities of generative AI for simulating people realistically – use of these capabilities in gaming and for other non-medical purposes
- Ways that generative AI provides the advantages of high-fidelity simulation without the disadvantages (guardrails)
- More realistic patients may stimulate renewed interest in PBL tutorials at a time when engagement in our own curriculum and elsewhere is lacking (Dawood, Rea, Decker, Kelley, & Cianciolo, 2021; Walling, Istas, Bonaminio, Paolo, Fontes, Davis, & Berardo, 2017; Kilgour, Grundy, & Monrouxe, 2016).

Generative AI-simulated patients offer higher fidelity patient simulation, with potentially greater control, than what is currently feasible in most medical PBL curricula. However, AI development can be time-consuming and expensive, and the technology is still evolving.

Research is needed to explore the impact of using generative AI to simulate patients in PBL tutorials to understand how (and when) to best implement this new technology.

## Methods

### Overview

This study used a between-groups, mixed-methods approach (Schifferdecker & Reed, 2009) to

1. form a comprehensive picture of medical student interactions with a generative AI-simulated patient in a PBL tutorial, as compared to interactions with multimedia patient case materials; and
2. triangulate on the impact these interactions had on learning. Two groups of students gathered patient history information from a generative AI-enabled, 3D-animated avatar (AI condition). Two other student groups gathered patient history information from a multimedia database using keyword searching (Electronic PBL Module [ePBLM] condition). We used descriptive observation to explore student interactions with both forms of the simulated patient, and we quantitatively compared students’ perceptions of their learning experience and recall of patient history information across conditions.

### Context/Setting

This study took place in March 2024 at a small, public, community-based medical school in the Midwestern U.S. (class size = 72), which was the first institution to implement its PBL curriculum using ePBLMs (Ryan & Koschmann, 1994). The 4-year curriculum follows a 2+2 design in which the first 2 years focus on preclinical, basic science instruction in a classroom setting with periodic clinical exposure, and the second 2 years focus on clinical clerkships and electives. The preclinical curriculum comprises spirally organized organ-system-based units; Year 1 instruction emphasizes anatomy, physiology, and histology, and Year 2 emphasizes pathophysiology and pharmacology. The curriculum is delivered using a hybrid PBL format (Lim, 2012) in which participation in PBL tutorials is required, and attendance at correlated lectures is recommended, but not mandatory.

PBL tutorials in our preclinical curriculum comprise small groups of 6-8 students and a facilitator who meet multiple times per week (3 times/week in Year 1, 2 times/week in Year 2) to exercise clinical reasoning with real-world patient cases, discuss understanding of the case content, identify basic science knowledge gaps (i.e., learning issues), and develop shared knowledge of the patient’s condition and its treatment (Koschmann, Glen, & Conlee, 1997; Cianciolo, Kidd, & Murray, 2016). Procedurally, students work collaboratively to “open” the case, which involves gathering patient information from history-taking, physical examination, laboratory studies, and imaging to understand the patient’s medical concerns, form a differential diagnosis, and identify learning issues to address with self-directed learning. Following self-directed learning, students “close” the case by discussing their self-directed learning findings, the implications of their new knowledge for understanding the patient case, and the patient’s care outcomes. The students comprising PBL tutorial groups change each unit, for a total of 94 unique groups assigned across both years of the preclinical curriculum.

### Participants

Participants comprised 4 groups of second-year medical students (N = 26, 37% of the Class of 2026. Among the 14 participants who provided demographics, 79% self-identified as women and 20% self-identified as belonging to a marginalized racial/ethnic group. Students’ participation occurred during the first 2 days of the final unit in the preclinical curriculum (Endocrinology, Reproduction, and Gastroenterology [ERG]), approximately 3 months prior to taking Step 1 of the USMLE Board Examination, and after nearly 2 years’ experience in our PBL (ePBLM) curriculum. Participants were a convenience sample recruited via word of mouth by fellow second-year medical students serving as co-investigators in this study. There were no inclusion/exclusion criteria. Participants were allowed to choose their PBL tutorial group with knowledge of who else was in their group. Groups were then assigned to the AI or ePBLM condition depending on investigator availability and preference.

This study was deemed non-human subjects research by the Springfield Committee on Research Involving Human Subjects (6 February 2024). The facilitator for all 4 groups, a faculty member from the investigator team—who is locally considered a master PBL tutor with 16 years of tutoring experience—described the study to participants and received their verbal consent to participate with audio-recording. Participants were offered pizza and drinks during their tutorial, but no other incentives were provided.

### Materials

*Patient Case – Randy Rhodes*. The patient case used in this study is a 54-year-old man—Randy Rhodes—who presents to the family medicine clinic with a rash on his arm. His diagnosis is Diabetes Mellitus Type 2. The case is designed to teach pathology, pharmacology, and other topics associated with this diagnosis (e.g., obesity and related metabolic syndromes) as one of 20 cases comprising the ERG PBL curriculum. Participants interacted with Randy Rhodes to gather patient history information in one of 2 ways, ePBLM or generative AI:

*ePBLM.* ePBLMs have been used to simulate patients in our PBL curriculum for over 30 years. Developed locally to replace the spiral-bound notebooks of comprehensive case information provided to students to role-play the patient, ePBLMs allow students to gather case information serially by using keyword searches to simulate history-taking, physical examination, testing/imaging, and procedures (Ryan & Koschmann, 1994). The ePBLM system returns answers to students’ search queries as realistically as possible using multimedia. For instance, it provides actual laboratory test or imaging results where possible, and it returns canned dialogue snippets in response to patient history queries (e.g., “I’ve had this rash for 3 months.”) as well as offering information about patient progression throughout their treatment. Figure 1 presents a screen capture of the ePBLM interface for the Randy Rhodes case.

**Figure 1.**
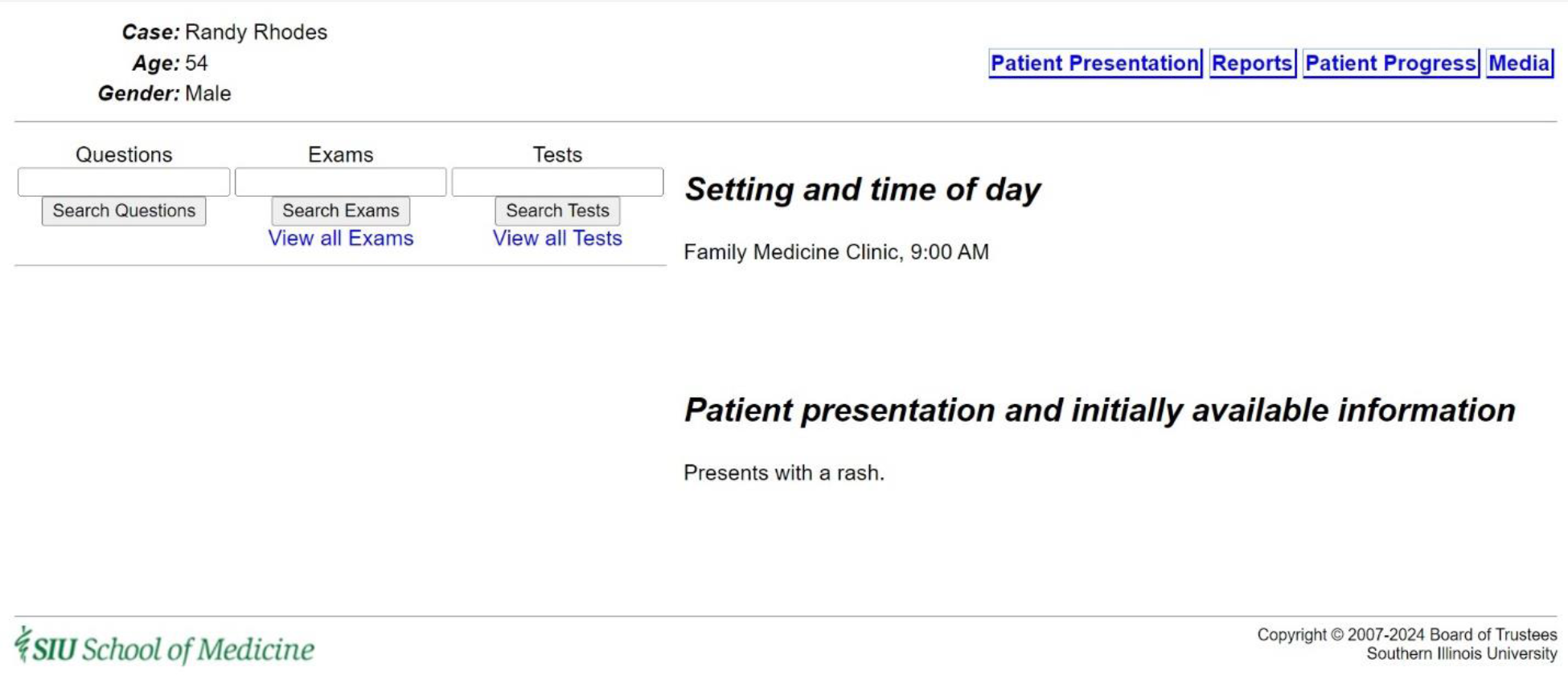
Screen Capture of the Randy Rhodes ePBLM

### Generative AI

Students gather patient history information from the AI-enabled patient via voice-to-voice interaction with a 3D-animated avatar situated in a clinical exam room environment created using Unreal Game Engine 5.0 and Metahuman (Figure 2). The content of the avatar’s responses to students’ history-taking questions—generated using the convai platform—is produced via prompts to a general large language model that have been engineered using the existing Randy Rhodes ePBLM database. The scope of the avatar’s responses is shaped by “guardrails” placed by the development team to optimize the case’s educational value. For instance, one guardrail prevents Randy Rhodes from revealing his diagnosis directly to students. Situated within a game engine, the AI also is capable of responding with different degrees of friendliness depending on how students interact with it. For example, the AI is trained to respond irritably to history questions laden with medical jargon and, if guardrails had not been implemented to prevent it, could storm out of the exam room.^1^ Interactions with the AI in this context may take forms other than questions; the avatar can respond to empathic statements and even tell jokes.

**Figure 2.**
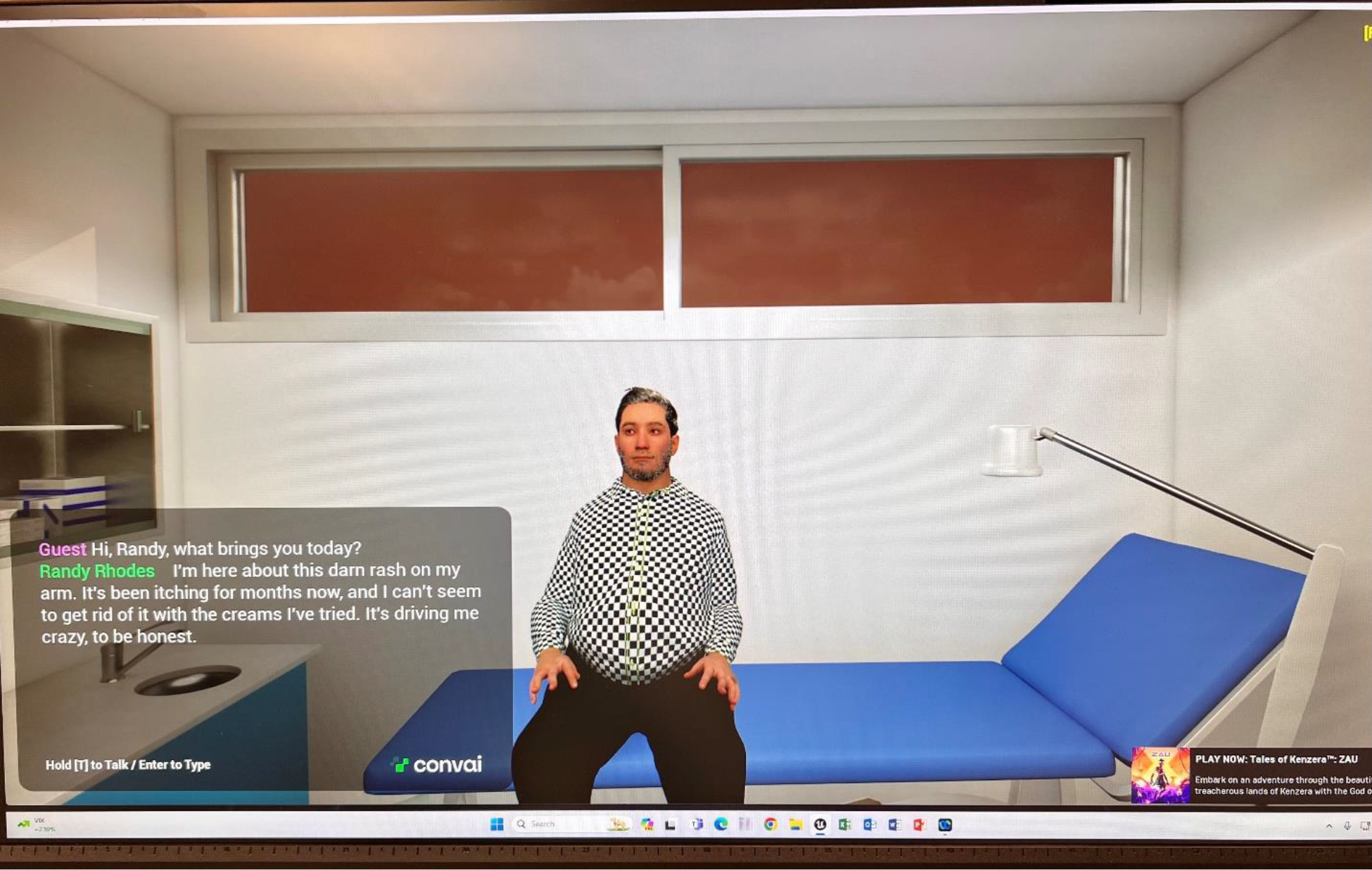
Screen Capture of Randy Rhodes, the Generative AI-enabled Patient

### Learner Perceptions Survey

This paper-based 8-item survey assessed participants’ perceptions of PBL tutorials using either ePBLM or AI. Participants used a scale of 1 to 5 to rate how positively they view several aspects of tutorial quality and impact (impact on improving clinical knowledge, clinical simulation accuracy, impact on clinical knowledge retention, enjoyability, stressfulness, allowance for teamwork, interestingness, impact on confidence) when either format is used. Both forms of the survey (ePBLM and AI) are presented in the supplemental appendix. A composite score for each form was created by summing the ratings across all 8 items (Item 5 was reverse coded). The possible range of scores was 8 to 40, representing how positively participants view their overall PBL tutorial experience using each format.

### Patient History Information Quiz

This paper-based quiz (supplemental appendix) consisted of 11 short-answer questions to assess participants’ knowledge of Randy Rhodes’s demographic information, medical history, and symptoms immediately following the PBL tutorial and 2 weeks later. Quizzes were graded by 2 student co-investigators masked to the condition (ePBLM or AI) and timing (immediate or delayed) when the quiz was completed. According to a faculty-verified key, each short answer could receive 0 points, ½ point, or 1 point, and the possible range of total scores was 0 to 11, representing overall recall of patient history information available in the Randy Rhodes case. Both co-investigators independently graded the first 5 quizzes to establish consistency with the rating criteria, after which 1 co-investigator graded the remaining 52 quizzes.

### Descriptive Observation Notes

Five co-investigators conducted descriptive observation (Spradley, 1980) of the audio-recorded tutorials, documenting their notes using a template created locally by a well-established medical education researcher (supplemental appendix). The template prompted observers to (1) document a reflexivity reflection; (2) specify (with time stamp) and describe noteworthy occurrences observed in the audio-recording; and (3) report in bracketed fashion their reactions to these noteworthy occurrences. The lead observer was a behavioral scientist with experience conducting observational research of PBL tutorials (Cianciolo, Kidd, & Murray, 2016). The other observers were first and second-year medical student co-investigators enrolled in our preclinical curriculum.

### Procedure

All 4 PBL tutorials took place in a small laboratory room arranged to mimic a typical PBL tutorial classroom (Figure 3). The room was equipped with tables, chairs, a whiteboard, and the tools needed to interact with the patient in each group: a keyboard and mouse to search the ePBLM database and a push-to-talk microphone to communicate verbally with the AI-enabled patient. The room also contained a computer monitor for presenting the simulated patient visually.

**Figure 3.**
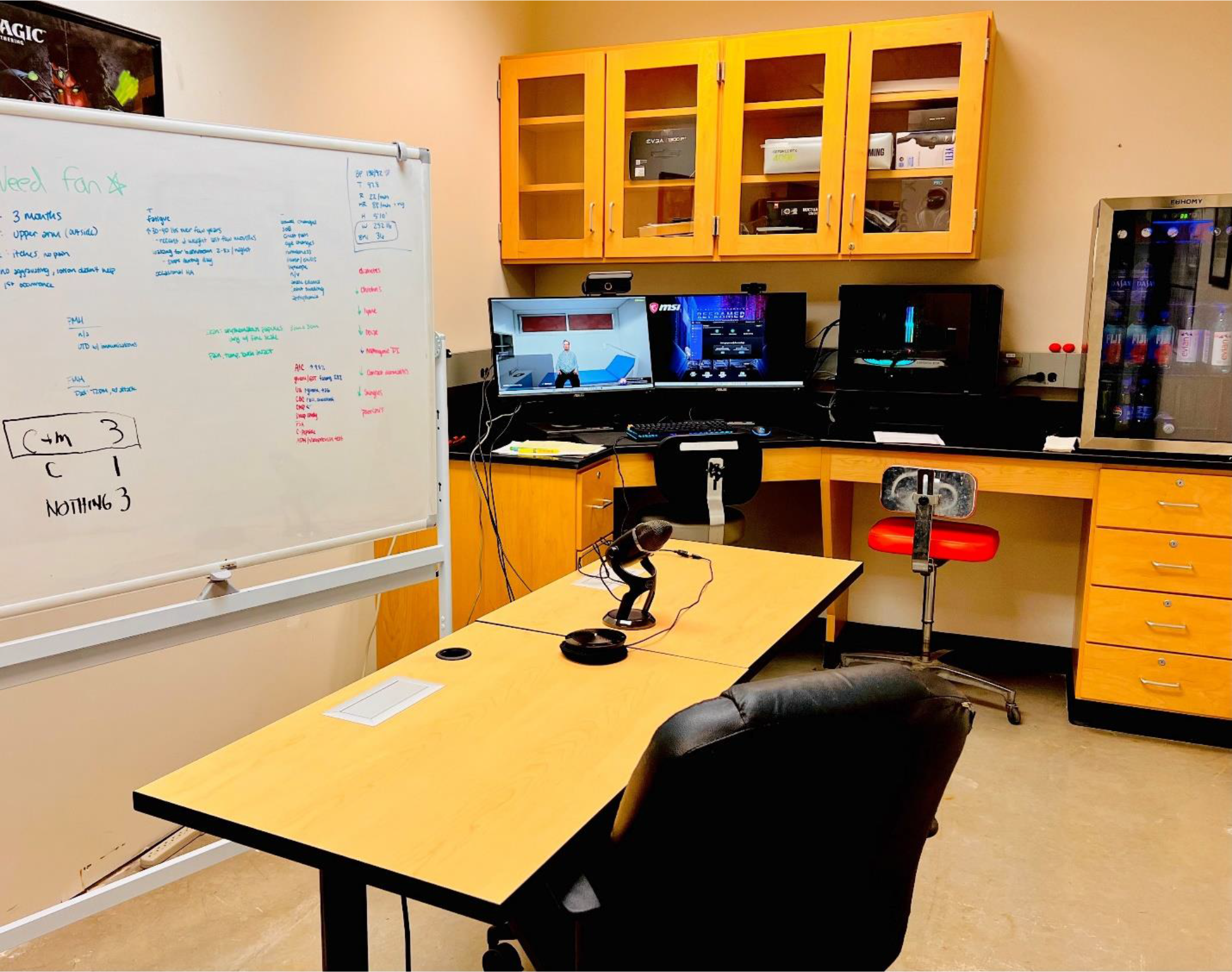
Photo of the Laboratory Room in Which the PBL Tutorials Took Place

After consent and before beginning their PBL tutorial, participants filled out the Learner Perceptions Survey – ePLBM. That is, participants rated their perceptions of ePBLM tutorials generally, based on their nearly 2 years’ experience in our PBL curriculum. Survey administration was proctored by 1 of 4 student co-investigators who attended the tutorials for this purpose as well as to answer participant questions, operate the push-to-talk microphone (AI groups), and manage the tutorial’s other technology, such as audio-recording. Audio-recording was accomplished via videoconferencing software (WebEx).

With minimal direction from the faculty facilitator, all 4 groups conducted a case opening, which ranged in duration from 36 to 75 minutes and consisted of history taking. After completing the case opening and before leaving the laboratory room, participants retook the Learner Perceptions Survey, this time rating their perceptions of PBL tutorials using the type of simulated patient they had just interacted with (ePBLM or AI). Participants also completed the Patient History Information Quiz.

Two weeks after the tutorials, participants individually retook the Patient History Information Quiz, proctored by a student co-investigator, in various campus locations of convenience for each participant. Student co-investigators graded the quizzes after all quizzes had been completed. Descriptive observation of the audio-recordings began after all 4 tutorials had occurred.

### Data Analysis

#### Quantitative analysis

Quantitative analyses were conducted using linear mixed models to control for within-group variance while examining the effect of condition (ePBLM vs. AI) on participants’ perceptions and recall of patient history information. P-values of .05 or smaller were deemed statistically significant. All analyses were performed using R version 4.3.2 (R Core Team 2023).

#### Qualitative analysis

Observation notes were analyzed inductively to characterize the interactions among students and the simulated patient in each group. For each of the 4 groups, the lead analyst compiled the independent observers’ notes into one set. These compiled notes were arranged chronologically to create a detailed, multi-perspective characterization of noteworthy events and associated bracketed reflections that occurred during each tutorial. To highlight the different perspectives at a glance, the lead analyst represented each observers’ notes using a different text color. Next, all qualitative analysts independently reviewed each set of compiled field notes to identify regularities in the interactions among people and case materials. Flagged as regularities were recurring interactional behaviors such as consistencies in patient questioning strategies or commonalities among occurrences of pauses or silence in group interactions. Analysis also flagged interactional behaviors potentially reflective of how students oriented to the patient case or each other (e.g., referring to the patient with personal pronouns or seeking input from peers in asking history-taking questions). Finally, all qualitative analysts collaboratively reviewed regularities and noteworthy interactional behaviors within and across participant groups to develop patterns in PBL behavior associated with type of simulated patient.

## Results

### Learner Perceptions Survey Data

Linear mixed modeling of the Learner Perceptions Survey data revealed a significant effect of time (pre- vs. post-tutorial) for the AI condition only (Table 1). Specifically, participants’ post-tutorial ratings of their overall PBL learning experience were, on average, 6 points higher than their pre-tutorial ratings if their tutorial involved an AI-enabled patient. In other words, students in the AI condition rated their present, AI-augmented PBL learning experience significantly higher overall than they rated their previous PBL learning experiences using ePBLMs. Students in the ePBLM condition perceived their present PBL learning experience to be approximately the same as their previous PBL learning experiences using the same technology.

**Table 1.** Linear Mixed Model of Learner Perceptions Survey Data.

| *Predictors* | **Survey total** |  |  |  |  |
| --- | --- | --- | --- | --- | --- |
|  | *Estimates* | *std. Error* | *CI* | *Statistic* | *p* |
| (Intercept) | 23.77 | 1.13 | 16.94 – 30.59 | 21.02 | **0.008** |
| Time: post | 0.23 | 0.98 | -1.80 – 2.26 | 0.23 | 0.817 |
| Condition: AI | -1.38 | 1.60 | -6.49 – 3.72 | -0.87 | 0.451 |
| timepost:conditionAI | 5.77 | 1.39 | 2.90 – 8.64 | 4.14 | **<0.001** |
| **Random Effects** |  |  |  |  |  |
| $\sigma^2$ | 6.30 | | | | |
| $\tau_{00}$ id | 10.00 | | | | |
| $\tau_{00}$ group | 0.00 | | | | |
| $\tau_{11}$ group.conditionAI | 0.00 | | | | |
| $\rho_{01}$ group | -0.98 | | | | |
| ICC | 0.61 |  |  |  |  |
| $N_{\text{group}}$ | 4 | | | | |
| $N_{\text{id}}$ | 26 | | | | |
| Observations | 52 |  |  |  |  |
| Marginal $R^2$ / Conditional $R^2$ | 0.241 / 0.707 | | | | |

The item-level data in Table 2 indicate that students’ post-tutorial perceptions of their learning experience in the AI condition were higher than their pre-tutorial perceptions almost across the board. Particularly noteworthy is the difference in perceptions of clinical simulation accuracy; on average, students who gathered patient history information from an AI-enabled patient rated the clinical accuracy of their present PBL tutorial 1.6 points higher (on a 5-point scale) than the clinical accuracy of their previous PBL tutorials using ePBLMs.

**Table 2.** Learner Perceptions Survey Item-Level Data – Mean (SD)

| Survey Item | Pre-Tutorial (N = 26) |  | Post-Tutorial (N = 26) |  |
| --- | --- | --- | --- | --- |
|  | AI (N = 13) | ePBLM (N = 13) | AI (N = 13) | ePBLM (N = 13) |
| Improves clinical knowledge | 3.8 (0.6) | 3.8 (0.9) | 4.2 (0.6) | 3.6 (0.7) |
| Simulates clinical experiences accurately | 2.6 (1.0) | 3.1 (1.0) | 4.2 (0.6) | 3.5 (0.9) |
| Helps clinical knowledge retention | 3.6 (1.0) | 3.6 (1.0) | 4.3 (0.6) | 3.5 (0.9) |
| Is enjoyable | 3.5 (0.8) | 3.7 (0.6) | 4.7 (0.5) | 3.8 (0.8) |
| Is stressful* | 4.1 (0.5) | 3.9 (0.5) | 3.8 (0.8) | 4.2 (0.6) |
| Allows for teamwork | 3.8 (0.4) | 4.0 (0.4) | 4.5 (0.7) | 4.0 (0.6) |
| Stimulates interest in learning the disease at hand | 3.4 (0.9) | 3.8 (0.7) | 4.4 (0.5) | 3.8 (0.8) |
| Improves confidence interacting with this chief complaint | 3.5 (1.0) | 3.8 (0.7) | 4.2 (0.8) | 3.5 (0.7) |
| TOTAL | 28.4 (0.4) | 29.8 (0.3) | 34.4 (0.3) | 30.0 (0.3) |

\*This item is reverse-coded such that a higher score represents lower perceived stress.

### Patient History Quiz Data

Linear mixed modeling of the Patient History Quiz data revealed a significant effect of time only (Table 3). Specifically, students’ recall of Randy Rhodes’s patient history information decreased significantly from immediate to delayed assessment, across both conditions (ePBLM and AI).

**Table 3.** Linear Mixed Model of Patient History Quiz Data.

| *Predictors* | **Quiz** |  |  |  |  |
| --- | --- | --- | --- | --- | --- |
|  | *Estimates* | *std. Error* | *CI* | *Statistic* | *p* |
| (Intercept) | 9.40 | 0.32 | 7.55 – 11.26 | 29.32 | **0.004** |
| Time: post | -1.46 | 0.29 | -2.05 – -0.87 | -5.07 | **<0.001** |
| Condition: AI | 0.69 | 0.41 | -1.09 – 2.48 | 1.71 | 0.233 |
| **Random Effects** |  |  |  |  |  |
| $\sigma^2$ | 1.08 | | | | |
| $\tau_{00}$ id | 0.50 | | | | |
| $\tau_{00}$ group | 0.00 | | | | |
| $\tau_{11}$ group.conditionAI | 0.00 | | | | |
| $\rho_{01}$ group | -1.00 | | | | |
| N<sub>group</sub> | 4 |  |  |  |  |
| N<sub>id</sub> | 26 |  |  |  |  |
| Observations | 52 |  |  |  |  |
| Marginal R<sup>2</sup> / Conditional R<sup>2</sup> | 0.382 / NA |  |  |  |  |

Immediate and delayed recall of patient history information slightly favored the AI condition (10.1 vs. 9.4 immediate recall for the AI and ePBLM conditions, respectively; 8.6 vs. 7.9 delayed recall for the AI and ePBLM conditions, respectively), however this difference was nonsignificant, and the rate of forgetting did not differ between the 2 conditions. Of note, immediate recall performance was near ceiling (maximum quiz score = 11 points) in both conditions, and delayed recall was also very high.

### Observation Data

Initial impressions from the observation of one group in the AI condition:

- The AI presented case content accurately, with an appropriate amount of information provided in response to students’ questions. The students appeared to find the avatar’s appearance, voice quality, and behavior plausible – with no language to suggest an aversive reaction to the patient representation (i.e., being in the uncanny valley).
- Although students used language suggestive of anthropomorphizing of the AI (e.g., gender pronouns, inferences about the AI’s emotional state), they appeared to orient to it as an augmented “question bank” for gathering patient history information. The format and style of their questions was consistent with how patient history information is gathered from the ePBLM (i.e., singularly focused on diagnosis) versus a more naturalistic conversation with the AI (i.e., diagnostically oriented, yet inclusive of rapport-building). Potential explanations for this include:
- `o` As rising third-years, the students were very experienced with using ePBLMs, and likely brought this framework for conducting PBL to the study.

♣ This frame may have been reinforced by using a patient case that is a required part of the curriculum versus an optional, experimental case.

♣ Further, the faculty facilitator did not offer participants a divergent perspective for how the AI might be used.

`o` Voice communication with the AI was mediated by a student co-investigator, who operated the push-to-talk microphone at students’ request. The additional layer between the students and the AI may have created a communication bottleneck that reduced the likelihood of naturalistic interactions.
“ Students were very engaged, as a group, in history taking with the AI-enabled Randy Rhodes.
`o` Students seemed to enjoy the novelty of the AI avatar, commenting on its appearance and reactions to what they said. The whole group seemed engaged in troubleshooting the AI’s unexpected behaviors (i.e., irritability).
`o` To communicate effectively with the AI, students had to frame their questions naturalistically instead of in terms of the keywords they were used to using. The whole group seemed engaged in troubleshooting what questions to ask.

## Discussion

Although using a generative AI-enabled patient simulation in a PBL tutorial had limited impact on recall of patient history information—which was near ceiling in both the ePBLM and AI conditions—students in the AI condition reported that the tutorial was more enjoyable and facilitative of teamwork than did students in the ePBLM condition. Descriptive observation of participants’ audio-recorded interactions in the PBL tutorials reinforced students’ self-reports and help to characterize what was enjoyable and engaging about taking a history from an artificially intelligent patient. Observation also provides insight into how to maximize the impact of generative AI-enabled patient simulations for clinical learning in the PBL context.

Results in the context of the PBL literature WRT patient simulation. Implications of results for optimizing AI implementation in PBL.

- Start implementation of AI early, perhaps in the first year, before an alternative framework can take hold.
- Provide students with an orientation to the AI agent to help them understand its capabilities (perhaps during the Year 1 orientation to PBL).
- Encourage “play” with or “discovery learning” of the AI’s capabilities.

Study limitations

- One case at one medical school (but more than one-third of the class)
- Study involved students very experienced with PBL and in the final unit of preclinical instruction preceding the Step 1 board examination
- Students were not randomly allocated to groups (although groups were randomly allocated to condition, and there did not appear to be systematic differences across condition in pre-existing social connections)
- Lack of video data to conduct more comprehensive observations
- Lack of a human-simulated patient to offer a comparison case (e.g., it is impossible to know whether students would orient to a human role-player as an augmented question bank)

Future applications of AI in PBL and elsewhere in the medical curriculum

- More diverse and varied patient interactions than traditional standardized patients while saving on time, scheduling, and training of standardized patients
- Potential for simulating telehealth visits and conducting standardized patient examinations
- Communication skills assessment (e.g., use of accessible language) through the adaptability of the AI to provide slightly different responses based on the phrasing of users’ questions
- Scalability of AI across different stages of medical education, from preclinical students learning to communicate and diagnose to clinical students learning treatment and management to residents and fellows learning to navigate complex or uncommon patient interactions

## Data Availability

All data produced in the present study are available upon reasonable request to the authors.

## Acknowledgements

We would like to thank Don Torry, PhD, for his guidance and continued focus related to research strategy; Debra L. Klamen, MD, MHPE, for her continued support for this project; and Sean McGinity BS, BA for his support to the quantitative data analysis. Finally, we would like to acknowledge the medical students from the Class of 2026 who participated in this study.

## Previous presentations

Earlier versions of this work were presented at the 2024 Central Group on Educational Affairs Spring Meeting, Milwaukee, Wisconsin, April 3-5, 2024 and at the SIU- SOM 14th Annual Teaching and Learning Symposium, Springfield, IL, April 11, 2024.

## Ethical approval

This project was deemed non-human subjects research by the Springfield Committee on Research Involving Human Subjects (6 February 2024).

## Supplemental Appendix

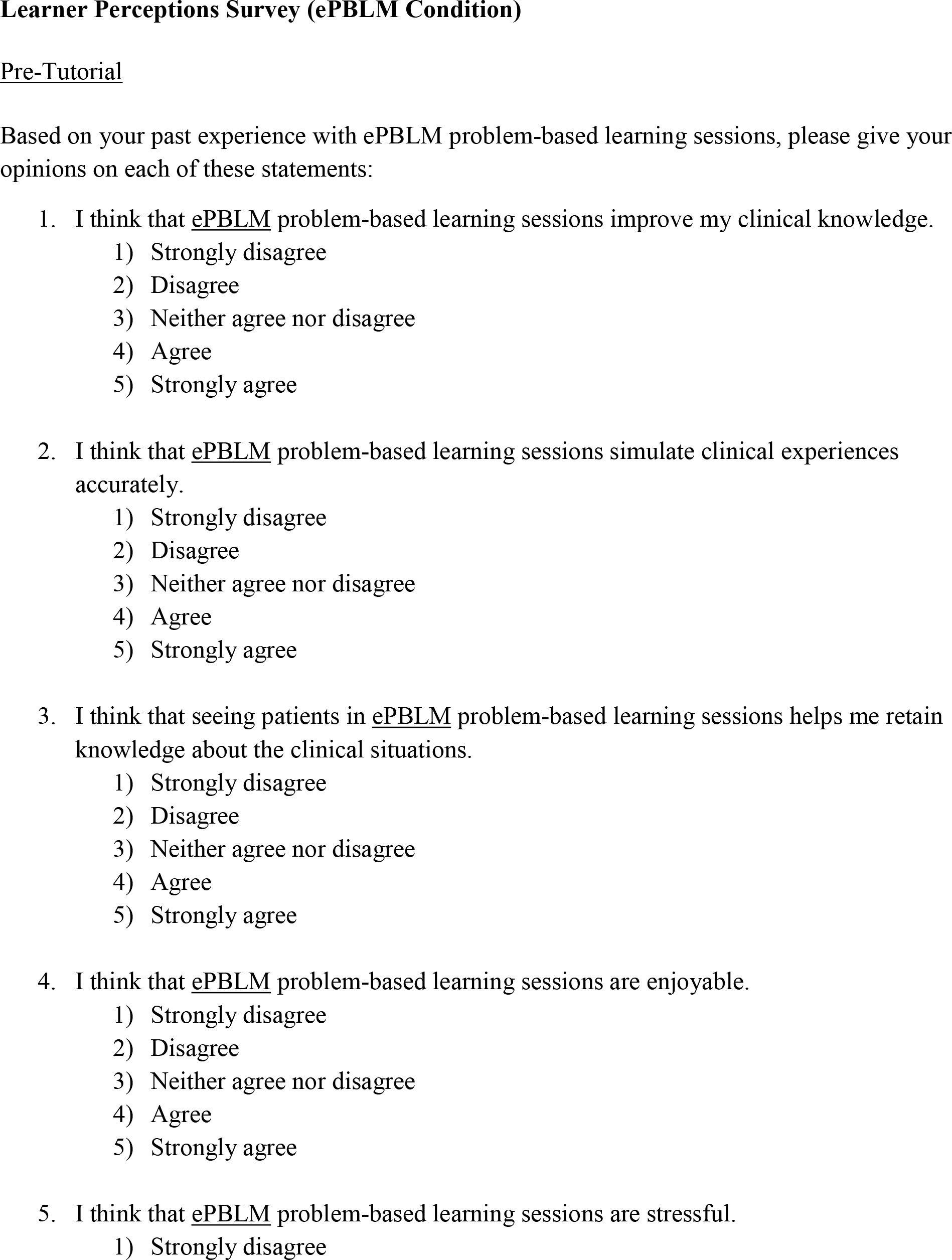
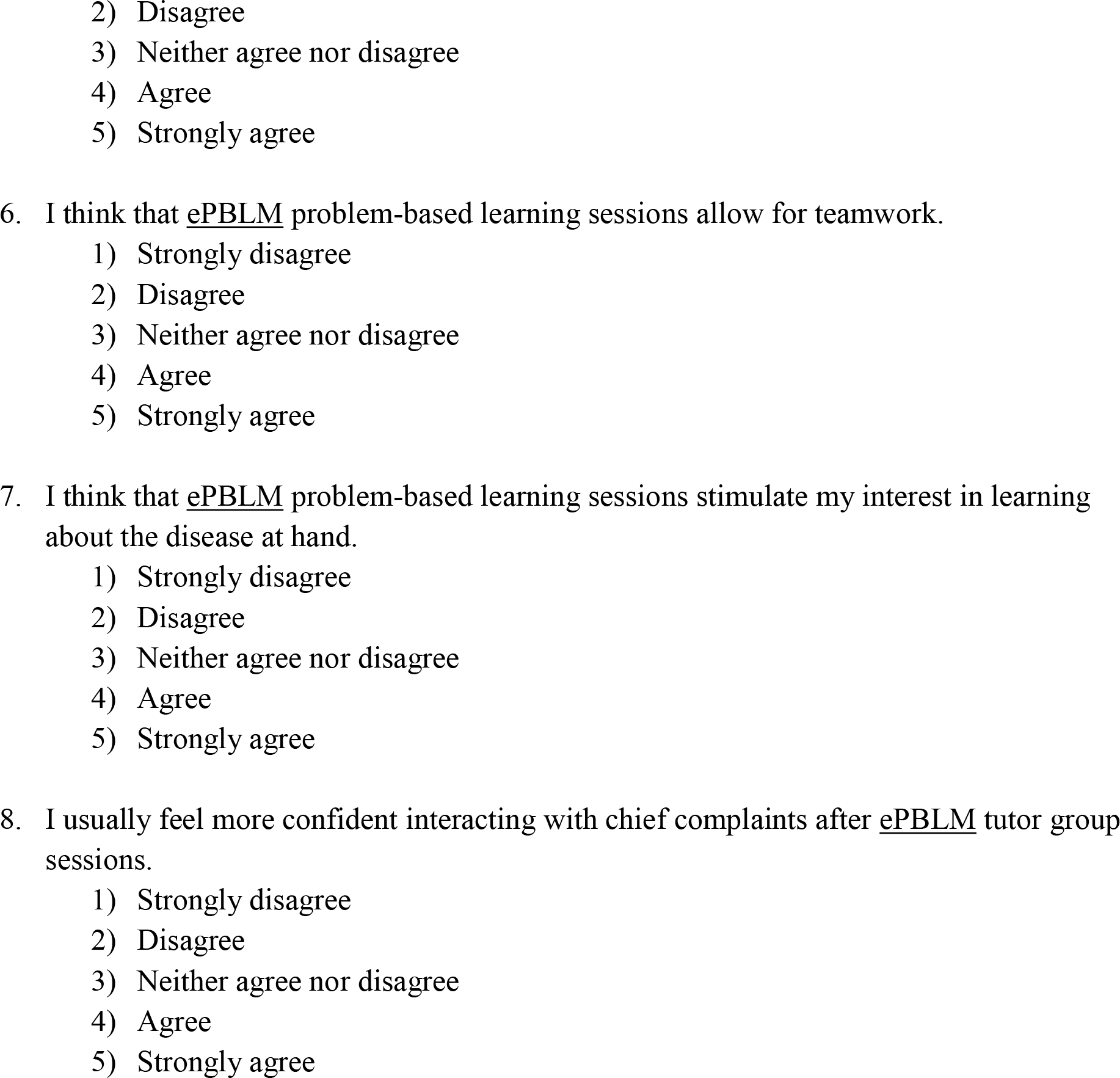
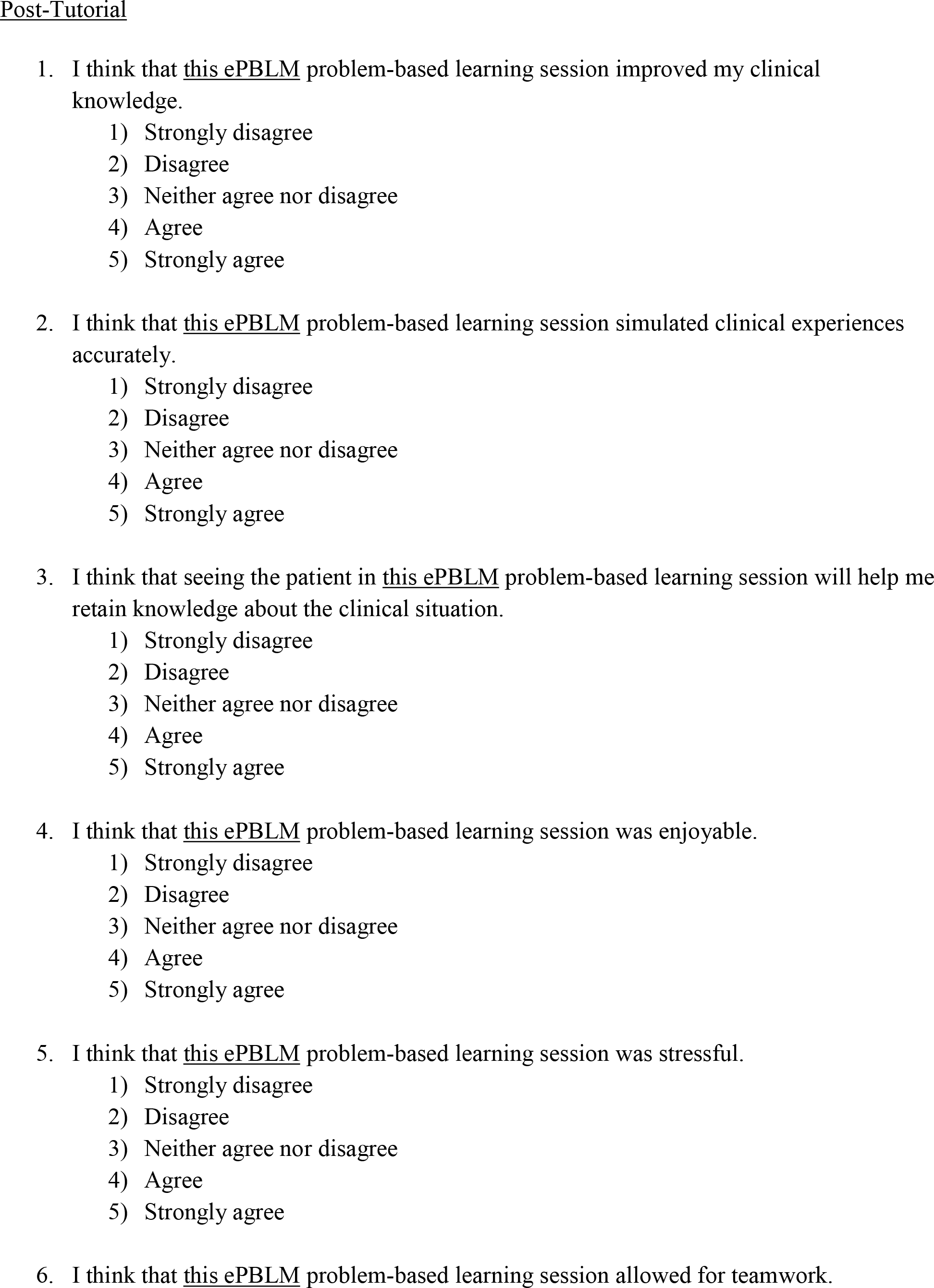
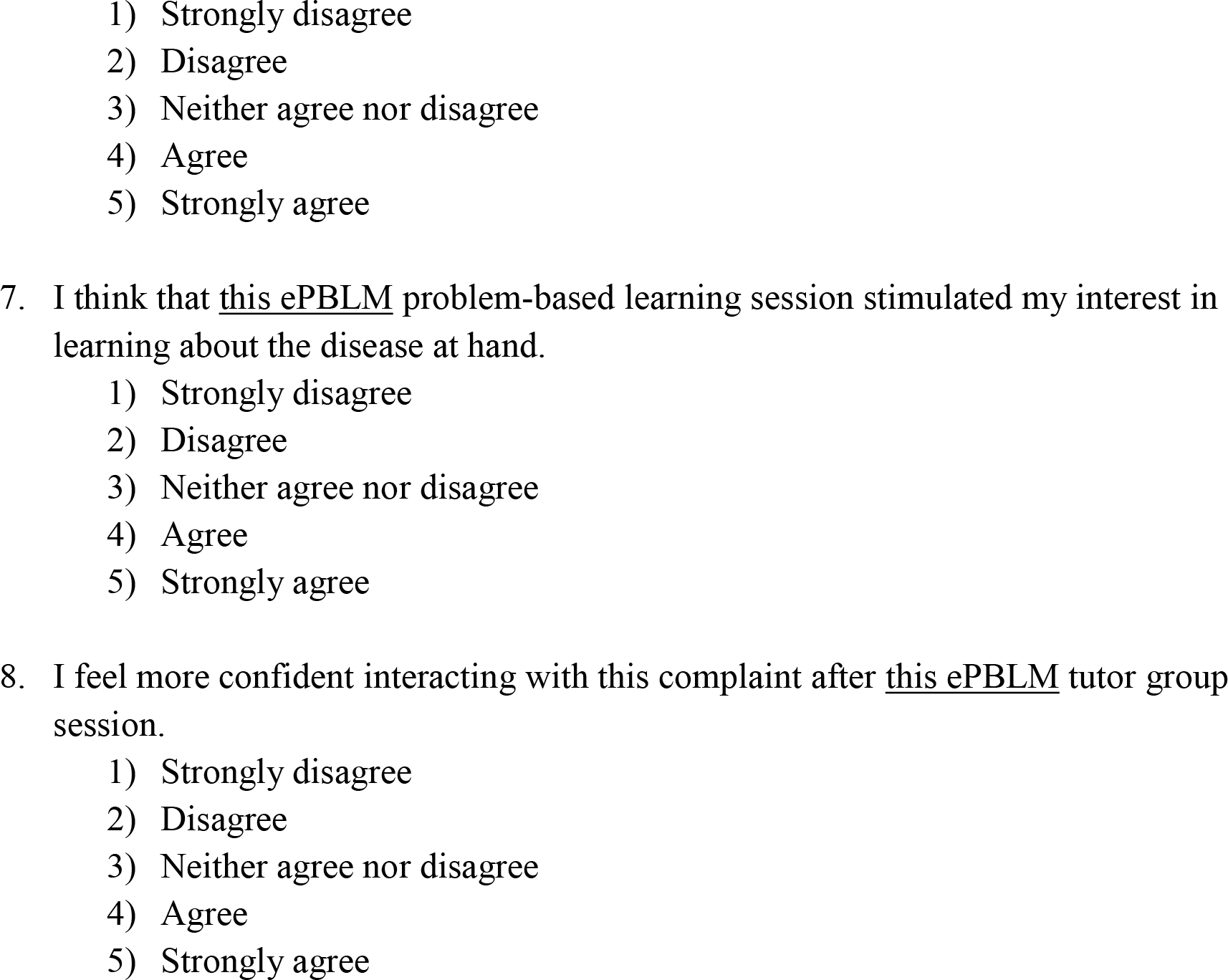
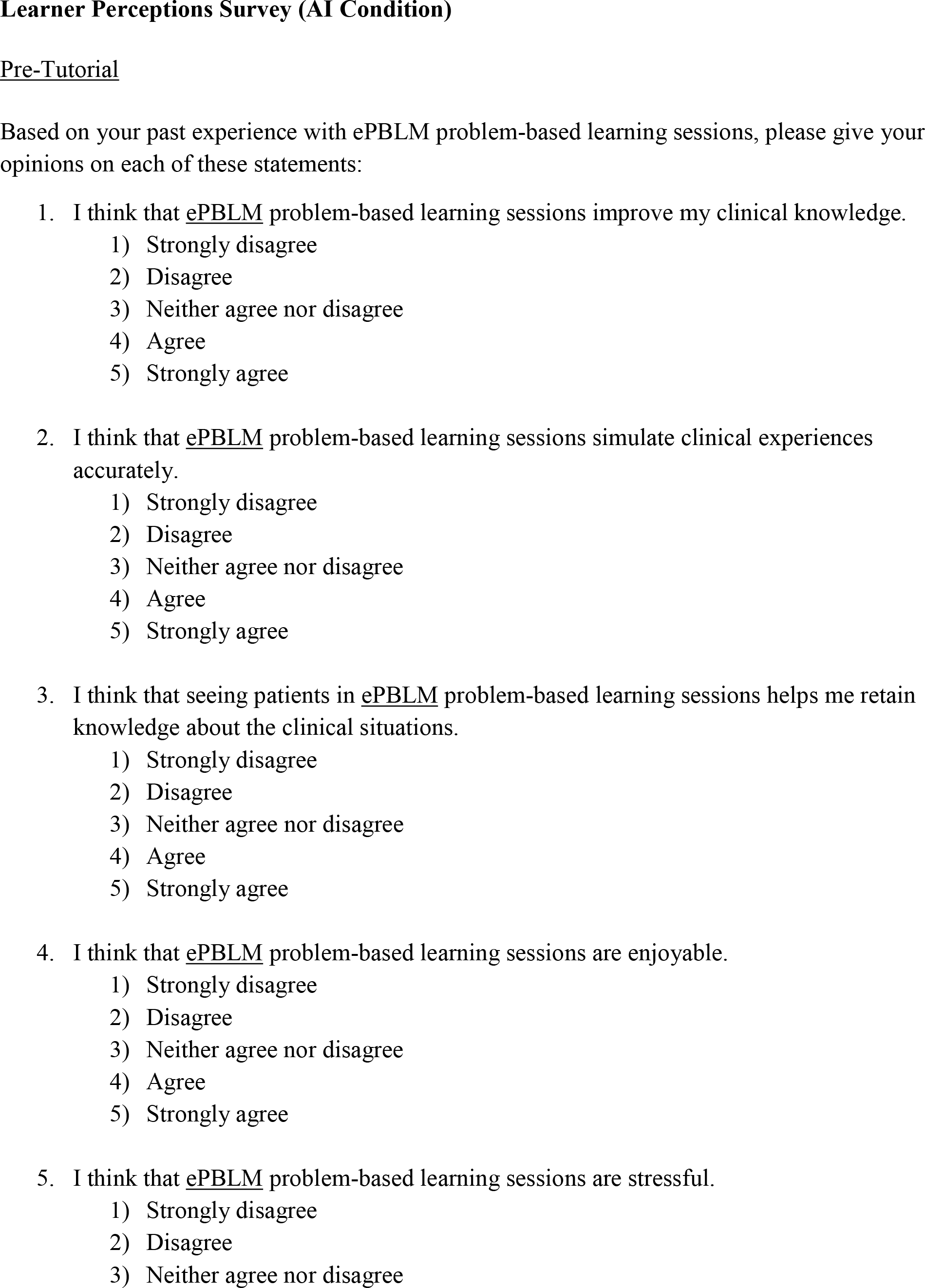
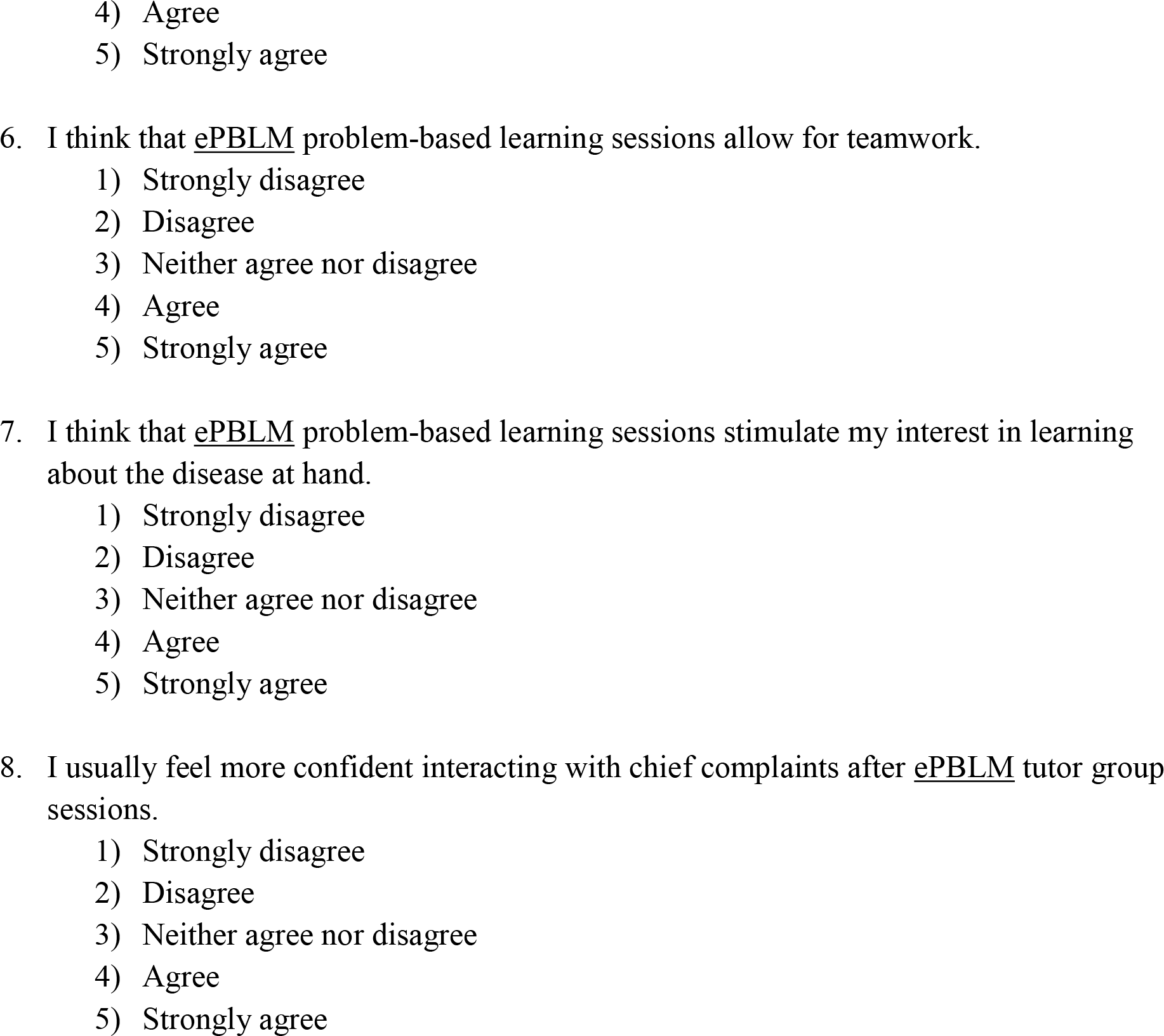
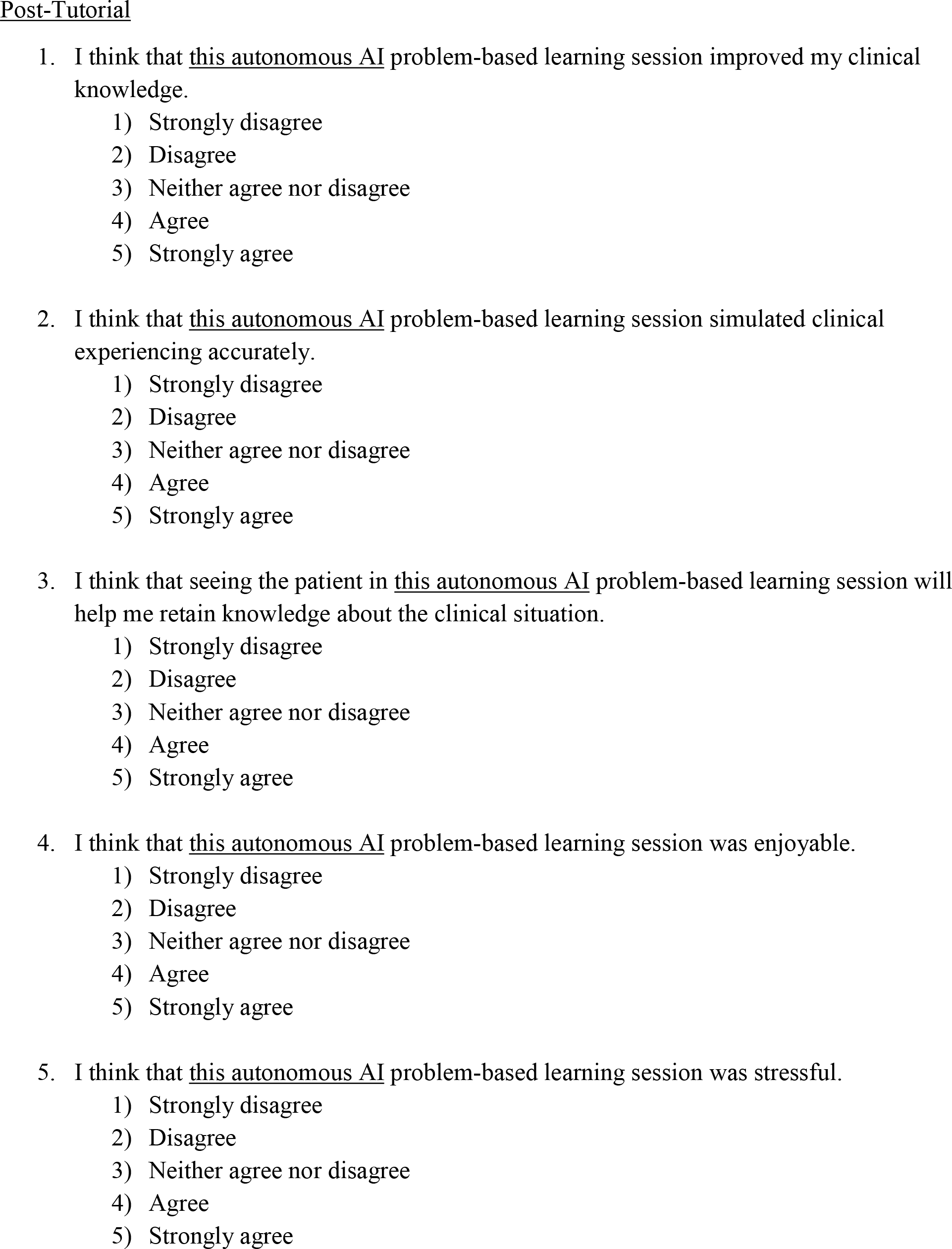
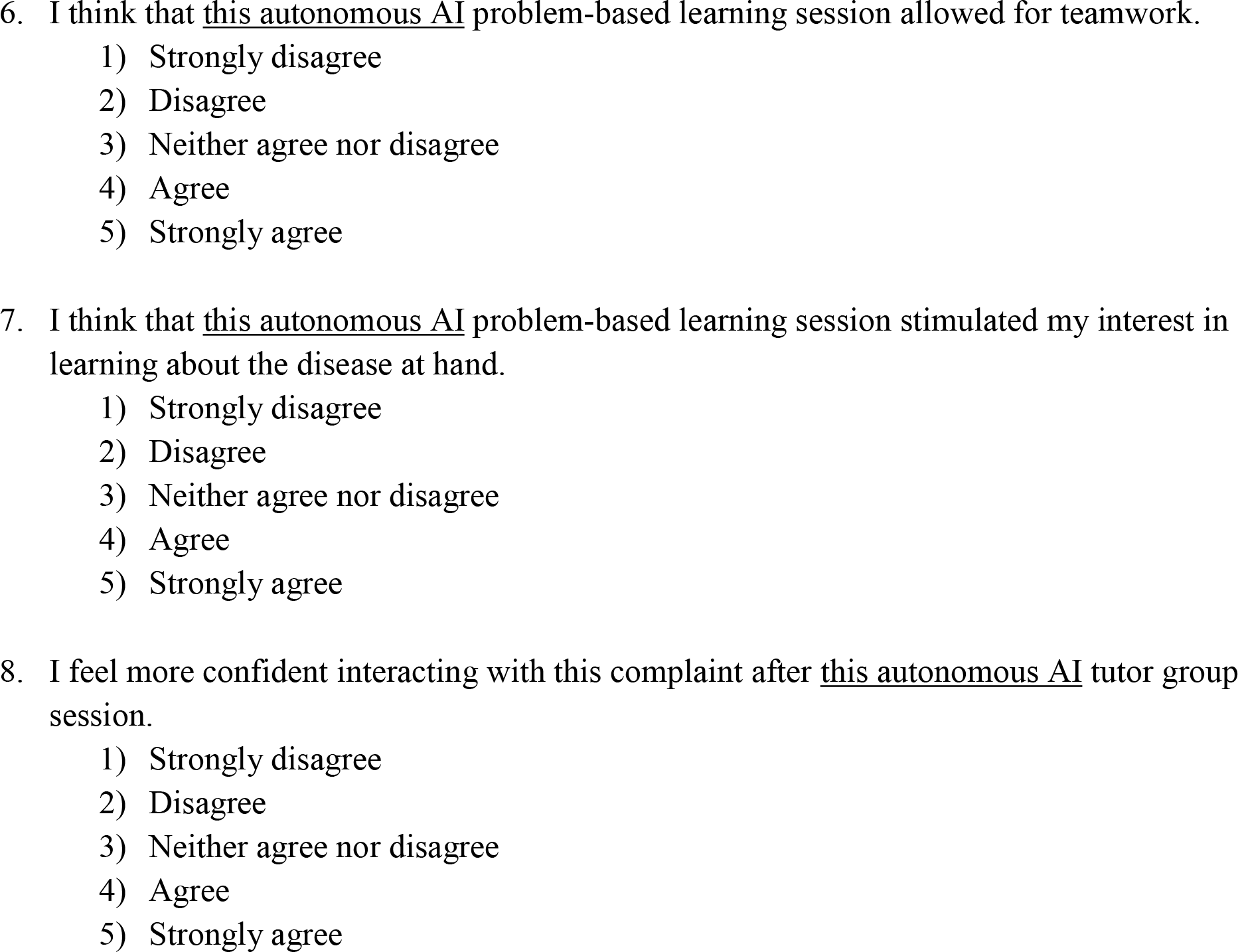
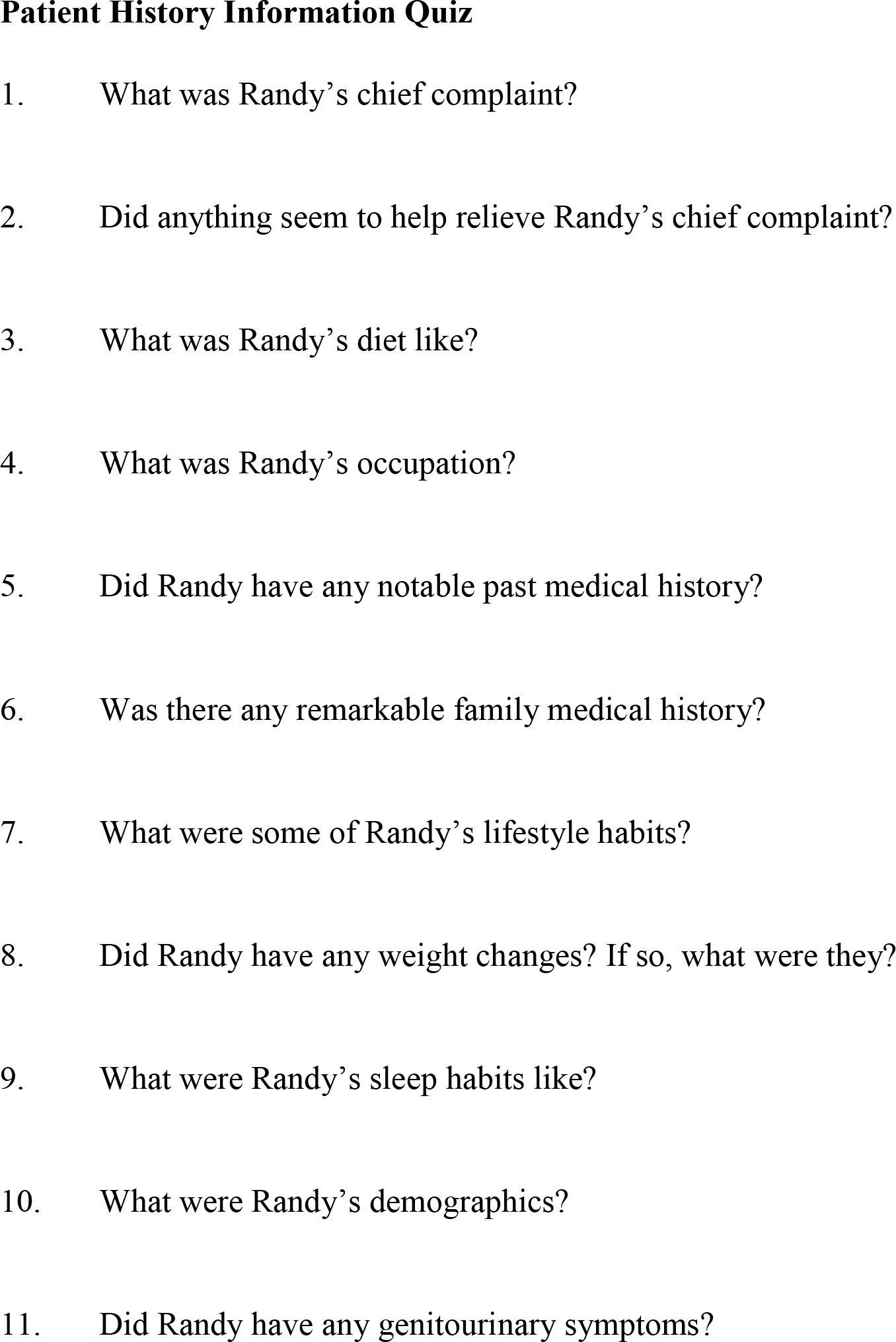

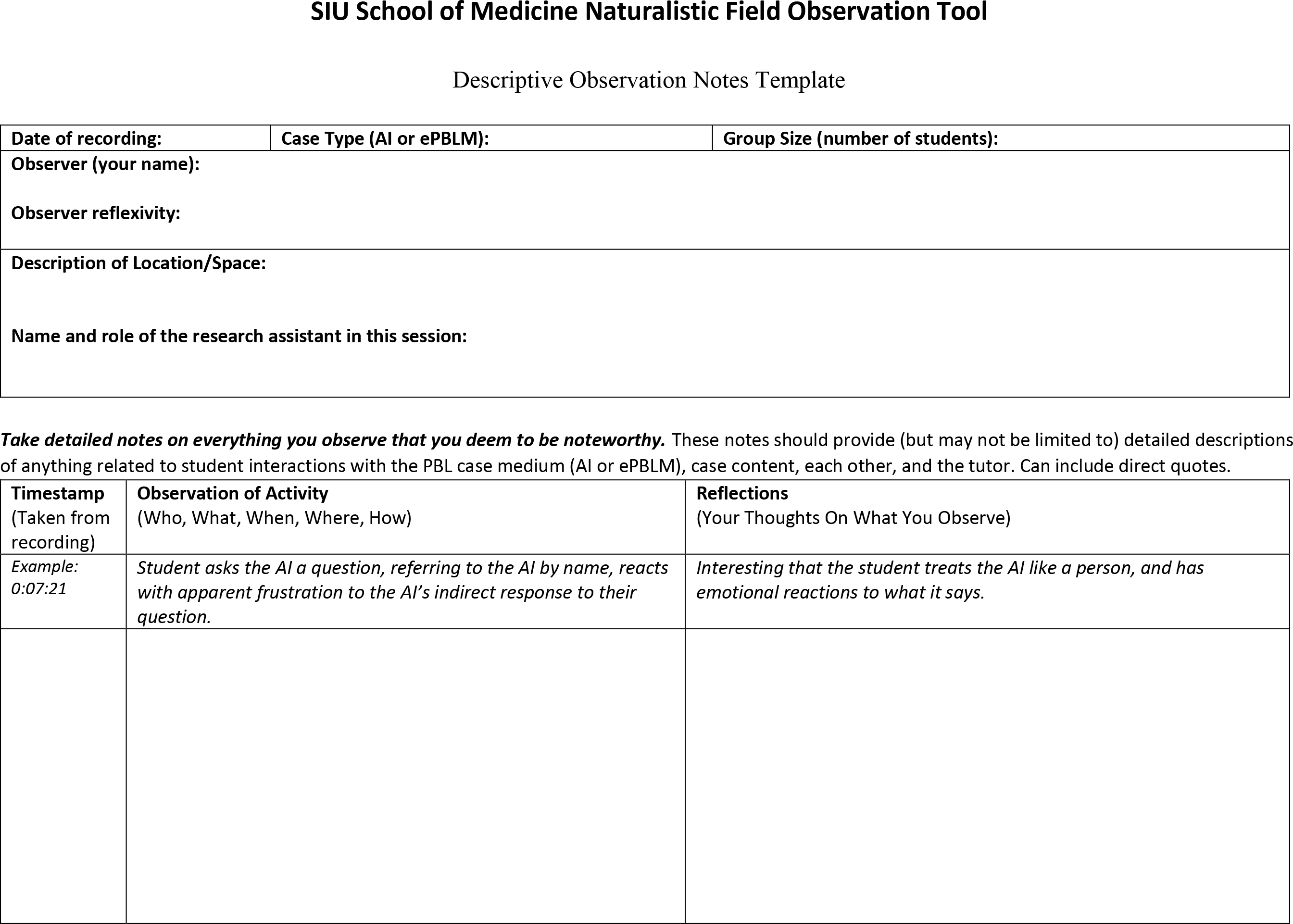

## Footnotes

1 Randy’s developers removed the door from the examination room such that storming out is no longer possible. Instead, the avatar will disengage from a problematic interaction and steer the conversation back toward the rash on his arm, which is his pressing concern and motivation for seeing a doctor.

## References

1. Wee LK, Kek MY, Sim MH. Crafting effective problems for problem-based learning. In Proceedings of the 3rd Asia-Pacific Conference on Problem-Based Learning. 2001 Jan 1.

2. Azer SA, Peterson R, Guerrero APS, Edgren G. Twelve tips for constructing problem-based learning cases. Med Teach. 2012;34:361–367.

3. Barrows HS, Tamblyn RM. Problem-based learning: an approach to medical education. Springer Publishing Company; 1980.

4. MacLeod A. Six ways problem-based learning cases can sabotage patient-centered medical education. Acad Med. 2011;86:818–825.

5. Cianciolo AT, Regehr G. Learning theory and educational intervention: producing meaningful evidence of impact through layered analysis. Acad Med. 2019;94(6):789–794.

6. Dawood O, Rea J, Decker N, Kelley T, Cianciolo AT. Problem-based learning about problem-based learning: lessons learned from a student-led initiative to improve tutor group interaction. Med Sci Educ. 2021;31:395–399.

7. Walling A, Istas K, Bonaminio GA, Paolo AM, Fontes JD, Davis N, Berardo BA. Medical student perspectives of active learning: a focus group study. Teach Learn Med. 2017;29(2):173–180.

8. Kilgour JM, Grundy L, Monrouxe LV. A rapid review of the factors affecting healthcare students’ satisfaction with small-group, active learning methods. Teach Learn Med. 2016;28(1):15–25.

9. Schifferdecker KE, Reed VA. Using mixed methods research in medical education: basic guidelines for researchers Med Educ. 2009;43(7):637–644.

10. Lim WK. Dysfunctional problem-based learning curricula: Resolving the problem. BMC Med Educ 2012;12:89.

11. Koschmann T, Glenn P, Conlee M. Analyzing the emergence of a learning issue in a problem-based learning meeting. Med Educ Online. 1997;2(1). 10.3402/meo.v2i.4290

12. Cianciolo AT, Kidd B, Murray S. Observational analysis of near-peer and faculty tutoring in problem-based learning groups. Med Educ. 2016;50(7):757–767.

13. Ryan C, Koschmann T. The Collaborative Learning Laboratory: a technology-enriched environment to support problem-based learning. Paper presented at the National Educational Computing Conference, 1994, Boston, MA. Available at: https://files.eric.ed.gov/fulltext/ED396678.pdf.

14. Spradley JP. Participant observation. Wadsworth: Thomson Learning. 1980.

